# Evaluation of the effects of transcranial direct current stimulation on the effectiveness of cognitive function rehabilitation using the RehaCom system in patients with schizophrenia: Study Protocol for a Randomized Controlled Trial

**DOI:** 10.64898/2026.04.01.26349996

**Authors:** Adam Wysokiński, Adrianna Szczakowska

**Author notes:** Corresponding author: Prof. Adam Wysokiński, MD PhD Medical University of Lodz, Department of Old Age Psychiatry and Psychotic Disorders Czechosłowacka 8/10 92-216 Łódź, Poland. Conflict of interest: Nothing to declare. Funding declaration: There was no external funding.

## Abstract

**Background:** Cognitive impairment is a core feature of schizophrenia and a major determinant of functional disability. Executive deficits affect approximately 85% of patients and are associated with reduced activity in the prefrontal cortex (hypofrontality). Current pharmacological treatments show limited efficacy in improving cognition, highlighting the need for alternative therapeutic approaches. Combining non-invasive brain stimulation with cognitive remediation may enhance neuroplasticity and improve cognitive outcomes.

**Methods:** This prospective, randomized, double-blind, sham-controlled, parallel-group superiority clinical trial. A total of 120 adults aged 18–65 years with clinically stable schizophrenia diagnosed according to DSM-5 criteria will be enrolled at a single clinical center. Participants will be randomly assigned in a 1:1 ratio to receive either active transcranial direct current stimulation (tDCS) targeting the dorsolateral prefrontal cortex followed by cognitive remediation therapy (CRT) using the RehaCom system, or sham stimulation followed by the same cognitive training. Assessments will be conducted at three time points: prior to the intervention (V1), immediately after the intervention (V2), and during the follow-up visit 8 weeks after the intervention (V3). The primary outcome is change in cognitive performance measured with the CANTAB battery. Secondary outcomes include symptom severity assessed with the PANSS, global clinical status (CGI-S), and neurophysiological changes measured by EEG. Written informed consent will be obtained from all participants, and the study has received ethics committee approval.

**Discussion:** This trial will evaluate whether tDCS administered prior to cognitive training enhances cognitive improvement compared with cognitive training alone. The findings may inform the development of more effective interventions targeting cognitive deficits in schizophrenia.

Trial registration ClinicalTrials.gov Identifier: NCT07273175. Registered on 25 November 2025.

## 1 Introduction

Schizophrenia is a chronic and severe psychiatric disorder that affects approximately 1% of the global population (Oertel, 2019; Zoupa, 2022). The disorder encompasses a heterogeneous range of clinical manifestations, including positive symptoms, such as delusions and hallucinations, negative symptoms, e.g., restricted affect, diminished emotional range, poverty of speech, reduced interests, decreased sense of purpose, and impaired social drive, as well as significant cognitive impairments (Zoupa, 2022; Buchanan, 2007).

It is estimated that cognitive deficits, occurring already in the prodromal phase and persisting over time, affect approximately 85% of individuals with schizophrenia, reducing their cognitive abilities by up to 90% compared to healthy individuals. Thus, they are a strong predictor of prognosis in schizophrenic individuals (Caponnetto, 2018; Green 2004). Cognitive dysfunctions have a greater impact on these patients’ adaptive and social functioning than other core symptoms, consequently limiting their ability to achieve personal autonomy, acquire new skills, obtain employment, and function effectively in society (Keefe et al., 2012; Velligan et al., 2000; Tomida et al., 2010). Among the most commonly impaired cognitive domains are learning, working and immediate memory, executive functions, verbal fluency, as well as attention and information processing (Saykin et al., 1991; Green, 2000). These deficits result from structural and functional abnormalities in the dorsolateral prefrontal cortex (dPFC) (Selemon, 1999; Pierri, 2001).

Cognitive deficits respond poorly to current pharmacotherapy (Dark, 2020; Keefe et al., 2012), which highlights the need to develop alternative approaches aimed at preventing disability in the course of schizophrenia. Recently, new interventions have emerged. They are based on neuroplasticity, i.e., a process involving structural and functional modifications of neuronal connections, aimed to train perceptual processes while simultaneously engaging the mechanisms of attention and working memory (Nitsche et al., 2012; Vinogradov et al., 2012). One such intervention is Cognitive Remediation Therapy (CRT), defined as an individualized therapy based on behavioral training through performance of specific exercises and tasks using a computer, paper and pencil, or through discussion (Radhakrishnan, 2016).

CRT is considered effective in the restoration of prefrontal functions and higher cognitive processes, and thus contributes to the improvement of daily functioning in individuals with such impairments (Caponnetto, 2018; McGurk et al., 2007; Wykes, 2011; Subramaniam et al., 2014; Fisher et al., 2010). This therapy also helps to reduce the number of hospitalizations compared to other rehabilitation strategies (Caponnetto, 2018). Moreover, neurocognitive rehabilitation techniques are well accepted by patients (Wykes et al., 2011). It has also been demonstrated that CRT leads to a significant increase in the activity of various brain regions—particularly within the frontal, prefrontal, occipital lobes, and the anterior cingulate cortex—during the performance of tasks related to working memory and executive functions (Isaac and Januel, 2016).

However, CRT requires many hours of repetitive and intensive exercises in order to induce significant changes. Moreover, cognitive training leads only to moderate improvement in cognitive functions, with an even smaller impact on daily functioning (McGurk SR, 2007). Therefore, combining cognitive rehabilitation with strategies that support neuroplasticity may not only produce yield greater and more durable improvements but also shorten the required training protocols (Jahshan, 2017). An example of such a strategy is neurostimulation, applied as a non-invasive tool for supporting cognitive functions (Dedoncker et al., 2016).

There are several neurostimulation methods with proven efficacy in improving cognitive functions (such as working memory and executive functions) in patients with schizophrenia. One of them is transcranial direct current stimulation (tDCS) (Hoy et al., 2014; Orlov et al., 2017a). It is a non-invasive technique that modulates neuronal activity by delivering a constant low-amplitude current (typically not exceeding 2 mA) to the scalp for a short period (not longer than 30 minutes) between two electrodes, i.e., the positive (anode) and the negative (cathode) (Nitsche et al., 2008; Elmasry, 2015; Abellaneda-Perez, 2020; Yamada, 2021a).

Due to its non-invasive nature and relatively low costs, as well as minimal risk associated with the very low current applied to the scalp, tDCS holds considerable clinical potential in the treatment of neuropsychiatric disorders associated with cognitive dysfunctions (Lally, 2013). Recent studies have shown that tDCS can enhance synaptic strength in neuronal pathways activated by cognitive training, thereby amplifying training effects and allowing for further expansion of cortical representation within the activated neural network (Elmasry, 2015; Miniussi & Vallar, 2011). Moreover, it has been demonstrated that in humans, tDCS can induce long-lasting increases in cortical excitability; just a few-minute-long stimulation can lead to changes in cortical excitability that persist for over an hour (Nitsche & Paulus, 2001), potentially enhancing the effectiveness of cognitive training administered immediately after stimulation. Based on these findings, several studies have investigated the potential benefits of combining these two intervention methods (Nienow, 2016; Orlov, 2017).

Our protocol is based on previous research showing that combining the two interventions, i.e., tDCS and cognitive training, may more effectively modulate brain regions responsible for cognitive functions in patients with schizophrenia. The primary aim of this study is to evaluate whether patients receiving DLPFC-targeted tDCS along with subsequent cognitive training using the RehaCom system achieve higher scores on the CANTAB test compared to those receiving sham stimulation and cognitive training with RehaCom. The authors hypothesize that tDCS combined with subsequent cognitive training can enhance neuroplasticity mechanisms, leading to improved performance on cognitive function assessments.

## 2 Methods

### 2.1 Roles and responsibilities

#### 2.1.1 Authorship and responsibilities

Principal Investigator: Prof. Adam Wysokiński, MD PhD, Department of Old Age Psychiatry and Psychotic Disorders, Medical University of Lodz-responsible for overall study oversight, protocol planning, study design, and data analysis. Study Coordinator: Adrianna Szczakowska MD, Department of Old Age Psychiatry and Psychotic Disorders, Medical University of Lodz– responsible for study logistics, participant recruitment, administration of tDCS interventions, and delivery of cognitive remediation therapy (CRT).

#### 2.1.2 Sponsor contact / Funding

Sponsor: Medical University of Lodz, ul. Kościuszki 4, 90-419 Łódź, Polska. The sponsor provides administrative and logistical support for the trial.

#### 2.1.4 Committees and data oversight

This trial does not have a separate steering committee, endpoint adjudication committee, or data monitoring board. The investigators are fully responsible for overseeing all aspects of the trial, including study conduct, data collection, quality control, safety monitoring, and reporting of results.

### 2.2 Study design

This prospective, randomized, double-blind, sham-controlled, parallel-group superiority experiment is designed to investigate the effect of transcranial direct current stimulation on the effectiveness of cognitive function rehabilitation using the RehaCom system in patients with schizophrenia. SPIRIT schedule of the study protocol is shown in Fig 1.

**Figure 1.**
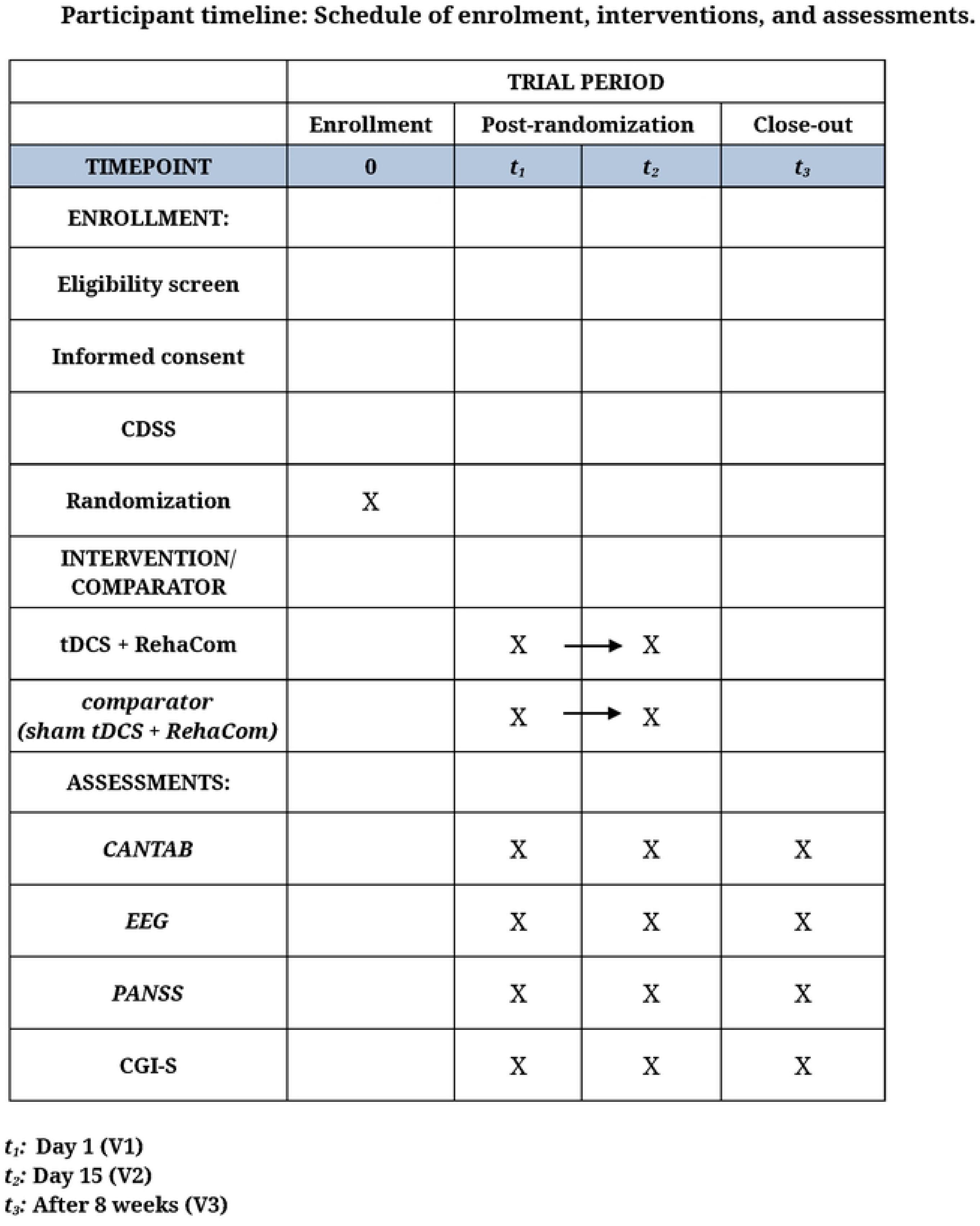
SPIRIT schedule.

The trial will be conducted at the Department of Old Age Psychiatry and Psychotic Disorders, Central Clinical Hospital, Łódź, Poland. All interventions and assessments will take place on site.

Eligible participants will be recruited and randomly allocated to one of two groups (1:1): individuals receiving active transcranial direct current stimulation (tDCS) that targets the dorsolateral prefrontal cortex (DLPFC) combined with RehaCom (Group 1), or participants undergoing sham tDCS combined with RehaCom, as a placebo control (Group 2). Randomization will be simple, with no stratification or adjustment for sex, age, or other participant characteristics.

A sham stimulation condition was selected as the comparator to control for placebo effects and to ensure adequate blinding. As tDCS is not an established standard treatment for cognitive impairment in schizophrenia, the use of a sham control is considered ethically justified. Importantly, all participants receive the same neurocognitive rehabilitation using the RehaCom system, ensuring that no participant is deprived of active therapeutic intervention.

Assessments will be conducted at baseline (visit 1; V1), after three weeks (visit 2; V2), and after eight weeks (visit 3; V3) using standardized instruments including CANTAB, EEG recording, the Positive and Negative Syndrome Scale (PANSS), and the Clinical Global Impression – Severity scale (CGI-S). Assessors will be trained and follow standardized procedures to ensure reliability. All forms and protocols for data collection are stored in the study database and are available upon request. The study design is presented in Fig 2.

**Figure 2.**
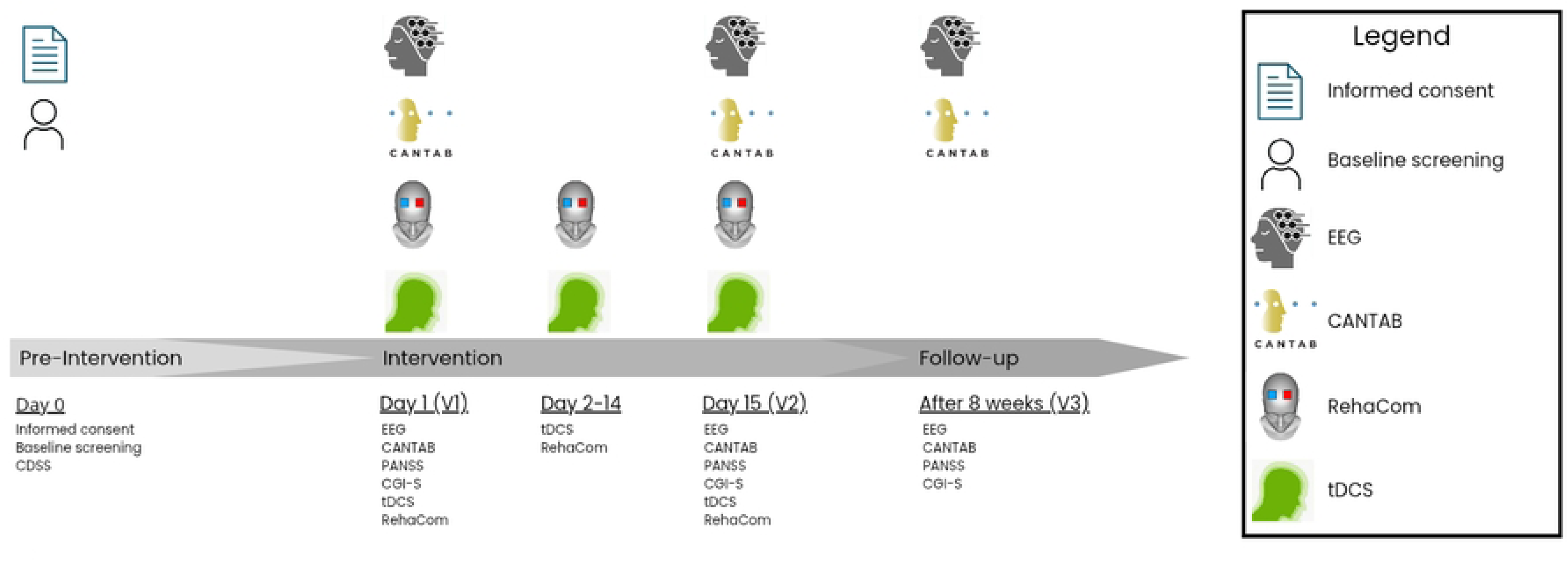
The study design.

Sessions are scheduled on five consecutive days (from Monday to Friday), with 24-hour intervals. Each participant will undergo 15 treatment sessions over a period of three weeks.

Attendance will be recorded daily, and participants missing more than 20% of sessions (i.e., more than 3 sessions) will be excluded from the per-protocol analysis. Reminders via phone will be used to improve adherence. Compliance with cognitive training will be monitored through RehaCom system logs, and research staff will supervise all sessions to ensure proper completion.

This study design allows for direct comparisons between active and sham stimulation conditions, and makes it possible to determine whether patients receiving active tDCS followed by neurocognitive training exhibit greater objective improvements in cognitive test performance compared to those undergoing sham stimulation combined with the same training protocol.

All participants and the researcher conducting the assessments will remain blinded during the study. As regards the implementation of blinding procedures, the authors adopted certain measures. Randomization will be conducted using a sham module in the battery-powered direct current stimulator (DC-Stimulator PLUS, neuroCare, Germany), which contains instructions with codes corresponding to real and sham stimulation. From these codes, another participant, not involved in data collection or analysis, will select 120 codes and record them in a random order. The study coordinator, responsible for assigning participants to interventions, will sequentially take a code for each newly recruited participant and enter it into the stimulator, which will automatically determine whether it is a sham or real stimulation code. Neither the participants, nor the personnel administering the stimulation, nor the outcome assessors will have access to the stimulator instructions, ensuring full allocation concealment. All will remain blinded for the entire duration of the study, and the codes will be revealed only after all study procedures are completed. This procedure ensures a completely random allocation of participants into the two study groups.

All participants, tDCS administrators, and outcome assessors will remain blinded to group allocation throughout the duration of the study. The unblinding procedure will normally be conducted after the completion of the study; however, in the event of a serious adverse event requiring knowledge of the intervention, the principal investigator—who has secure access to the allocation key—will be consulted, and the reason for unblinding will be documented.

The study protocol has been registered link to the study at ClinicalTrials.gov (Identifier: NCT07273175, <u>link to the study</u>) on 25 November 2025. The study started on 1 October 2023. Recruitment is ongoing (enrolling by invitation) and is expected to be completed before December 2026. Data collection for the primary outcome measures is anticipated to be completed by 3 December 2026. The overall study completion is expected on 3 December 2027, and the study results are anticipated thereafter.

Trial conduct will be regularly monitored by members of the research team, who will assess compliance with the study protocol, accuracy of data collection, and documentation of adverse events. Monitoring will include weekly reviews of recruitment progress, adherence to the visit schedule, and compliance with the intervention protocol. Patients and members of the public were not involved in the design, conduct, reporting, or dissemination of this study.

Any important protocol modifications will be submitted to the Ethics Committee for approval prior to implementation. All relevant members of the research team will be informed of approved changes, and trial registries or relevant authorities will be updated if applicable.

### 2.3 Participants

#### 2.3.1 Eligibility criteria

Participants will be adults, aged between 18 and 65 years, with schizophrenia classified based on the criteria outlined in the DSM-5 (Diagnostic and Statistical Manual of Mental Disorders, Fifth Edition). All participants will be required to provide informed consent for the tDCS treatment and for the use of collected data for research purposes. Patients must be mentally stable; they may receive pharmacological treatment.

#### 2.3.2 Exclusion criteria

The study will exclude individuals with contraindications to tDCS, including implanted electronic devices, metal implants in the head, a history of seizures, or contact allergy to materials used for tDCS stimulation (electrodes). Other exclusion criteria will be pregnancy, head injury with loss of consciousness or neurosurgery performed within the previous six months, history of stroke, aneurysm, brain tumor, or other conditions causing increased intracranial pressure, migraines, abuse of active substances, currently received electroconvulsive therapy (ECT). These exclusions aim to minimize confounding variables and ensure participant safety. The study will be discontinued in any of the following conditions occur: (1) urgent medical issues; (2) serious adverse events and side effects; or (3) unwillingness to continue participation in the trial.

#### 2.3.3 Recruitment and screening

Participants will be recruited from the Department of Psychiatry at a university hospital which includes an inpatient psychiatric ward and a day hospital unit, as well as from non-hospital settings (outpatient care facilities). Potential participants will be provided with both oral and written information regarding the study procedures and its potential benefits and risks. Those willing to participate will first complete a screening questionnaire aimed to assess basic eligibility criteria, such as age and presence of exclusionary conditions. Individuals meeting the initial criteria will be invited to an in-person screening appointment during which eligibility will be confirmed through structured interviews and validated behavioral assessments. The Calgary Depression Scale for Schizophrenia (CDSS) will be applied to rule out significant depressive symptoms that could affect the severity of cognitive dysfunction in study participants.

Recruitment progress will be monitored weekly to ensure the target sample size is reached within the planned timeline.

#### 2.3.4 Ethical approval and informed consent

Written informed consent was obtained from all participants prior to their enrollment in the study by trained study personnel. Participants were informed about the study objectives, procedures, potential risks and benefits, their right to withdraw at any time without consequences, and they provided consent for the collection, storage, and use of their study data for research purposes. The study will be conducted in accordance with the Declaration of Helsinki of 1995 (as revised in Edinburgh in 2000). The research was submitted for approval to the Ethics Committee of the Medical University of Łódź and initially approved under decision number RNN/244/19/KE on 09/04/2019. A subsequent approval for protocol amendment, including the addition of a new investigator, was granted under decision number KE/987/23 on 12/12/2023.

#### 2.3.5 Sample size

A final target of 120 participants will be equally divided and assigned to two study groups, i.e., a sham stimulation group and an active tDCS stimulation group. No *a priori* power analysis was performed, which results from the nature of the tool used to measure cognitive performance, i.e., CANTAB, a computerized battery of neurocognitive tests (described in details in the following sections). CANTAB gives very detailed results but no single performance score. Therefore, it is not possible to establish an expected improvement or calculate the required sample size. However, the authors will perform *post hoc* analysis and present effect sizes for all major outcomes.

### 2.4 Intervention

Participants will receive tDCS targeting the DLPFC, which was selected due to its key involvement in executive cognitive functions. These include working memory, planning, decision-making, inhibitory control, and the regulation of attention, all of which are relevant for modulating cognitive dysfunction in schizophrenia.

The tDCS will be administered using a battery-powered direct current stimulator (DC-Stimulator PLUS, neuroCare, Germany) through two saline-soaked sponge electrodes (5 cm × 7 cm). According to the International 10–20 system, the anodal electrodes will be positioned over the left DLPFC on the sites corresponding to F3, whereas the cathodal electrode will be placed above the right DLPFC, corresponding to F4. The electrode locations were verified using a computational brain model. All sessions will be conducted over five consecutive days (from Monday to Friday), with a 24-hour interval between sessions. Each participant will complete a total of 15 treatment sessions within a three-week period. Stimulation will be administered using the fixed parameters according to the established literature. For the active tDCS condition, a direct current of 2.0 mA will be applied (corresponding to a total charge of 2.4 C, a current density of 0.57 A/m^2^, and a charge density of 685.7 C/m^2^), for 20 minutes with ramp-up and ramp-down periods of 20 seconds each. For sham tDCS, the stimulation will utilize the same active tDCS arrangement, with an intensity of 2.0 mA. However, the current will be applied only for the 20 s ramp-up phase at the beginning and the 20 s ramp-down phase at the end of the stimulation. This protocol preserves blinding by mimicking the initial somatosensory sensations typically associated with active tDCS.

The selection of these parameters is grounded in prior evidence which demonstrates their functional specificity and efficacy in modulating cognitive processes in individuals diagnosed with schizophrenia.

All sessions will be conducted by trained personnel, and adverse events will be regularly monitored throughout the study.

Immediately after each tDCS stimulation, patients will undergo a 30-minute cognitive rehabilitation training session using the RehaCom computer system (HASOMED GmbH, Germany). Previous studies show that the time for which changes in neuronal excitability remain are proportional to the duration of stimulation and current used (Nitsche and Paulus, 2000). Based on these data, the authors assume that following a 20-minute tDCS session, the after-effects should last for at least 30 min (and hence the duration of neurocognitive training will be set at 30 min per session). The following programs will be used each week: day 1 (Monday), 3 (Wednesday) and 5 (Friday): EINK – shopping (different memory functions and selective attention), SUSA – selection of items (sustained attention), WOME – card games (working memory); day 2 (Tuesday) and 4 (Thursday): GEAU – maintaining the set vehicle speed (divided attention), LODE – creating logical sequences (logical reasoning); WOME – card games (working memory). During every session, each program will be applied for ten minutes, using a 17-inch laptop and a dedicated control panel. The following parameters will be set for SUSA: level up 85% and level down 70%; for LODE: upper threshold 90%, bottom threshold 60%, number of tasks per level 10; for WOME: number of tasks 10, repetitions 1, card display time 2000 ms; for GEAU: upper threshold 95%, bottom threshold 80%; for EINK: upper threshold 90%, bottom threshold 80%, repetitions 1, max time 90/300 s.

### 2.5 Outcome measures

The primary outcome of the study is the evaluation of the effect of tDCS intervention on the effectiveness of cognitive function rehabilitation using the RehaCom system in patients with schizophrenia. The scope of these changes will be assessed using the Cambridge Neuropsychological Test Automated Battery (CANTAB) test (Cambridge Cognition, UK). CANTAB will be administered at three time points: before the initiation of the stimulation cycle (V1), immediately after the completion of the three-week rehabilitation program (V2), and eight weeks later (V3) (see Fig 2). The CANTAB scores will be determined using the software. The results will present the parameters that have changed (improve/deteriorate) during the period of cognitive stimulation and rehabilitation. These measures aim to capture both the acute and cumulative effects of tDCS. The following CANTAB tests will be used:

1. DMS (Delayed Matching to Sample) – assesses both simultaneous visual matching ability and short-term visual recognition memory, for non-verbalisable patterns; outcome measures include latency (the participant’s speed of response), the number of correct patterns selected and a statistical measure giving the probability of an error after a correct or incorrect response for short-term visual memory. The duration of the test is seven minutes.
2. ERT (Emotion Recognition Task) – measures the ability to identify six basic emotions (sadness, happiness, fear, anger, disgust or surprise) in facial expressions. Computer-morphed images derived from the facial features of real individuals, each showing a specific emotion, are displayed on the screen, one at a time. Each face is displayed for 200ms and then immediately covered up. The outcome measures for ERT cover percentage and number of correct or incorrect and overall response latencies. The duration of the task is nine minutes.
3. MTT (Multitasking Test; a test of the participant’s ability to manage conflicting information provided by the direction of an arrow and its location on the screen, and the ability to ignore task-irrelevant information; outcome measures include response latencies and error scores that reflect the participant’s ability to manage multitasking and the interference of incongruent task-irrelevant information on task performance (i.e., a Stroop-like effect) for managing conflicting information. The test lasts eight minutes.
4. OTS (One Touch Stockings of Cambridge) – a test of executive function, based on the Tower of Hanoi test. It assesses both the spatial planning and the working memory subdomains. Outcome measures include the number of problems solved on first choice, mean choices to correct, mean latency (speed of response) to first choice and mean latency to correct for planning and working memory. The duration is ten minutes.
5. PAL (Paired Associates Learning) – outcome measures include errors made by the participant, the number of trials required to locate the pattern(s) correctly, memory scores and stages completed; for visual memory and new learning. The test lasts eight minutes.
6. RTI (Reaction Time) – assessments of motor and mental response speeds, as well as measures of movement time, reaction time, response accuracy and impulsivity; outcome measures are divided into reaction time and movement time for both the simple and five-choice variants for motor and mental response speeds, movement time, reaction time, response accuracy and impulsivity. The test lasts four minutes.
7. RVP (Rapid Visual Information Processing) – outcome measures cover latency (speed of response), probability of false alarms and sensitivity for sustained attention. The duration is nine minutes.
8. SWM (Spatial Working Memory) – requires retention and manipulation of visuospatial information; it also has notable executive function demands and provides a measure of strategy as well as working memory errors; outcome measures include errors (selecting boxes that have already been found to be empty and revisiting boxes which have already been found to contain a token) and strategy for measure of strategy as well as working memory errors. The duration of the task is four minutes.

Secondary outcomes will focus on the neural mechanisms underlying these behavioral changes. Resting EEG will be recorded at the same time points as the CANTAB battery to capture neurophysiological after-effects (Fig 2).

During V1, V2 and V3, the PANSS will be used to assess the severity of schizophrenia symptoms. It will include a positive symptom scale and a negative symptom scale, each containing seven items, as well as a general psychopathology scale with 16 items, i.e., 30 items in total. Each item on the PANSS will have a definition and a specific seven-point operational rating scale. The PANSS will be used to assess psychotic symptoms from different perspectives. At the same time points (V1, V2, and V3), the CGI-S will be used to evaluate both the severity of a patient’s condition and the efficacy of treatment. These measures will provide a broader understanding of individual differences among participants and options of potential modulation by tDCS.

Finally, follow-up assessments conducted eight weeks after the completion of the cognitive rehabilitation program will evaluate whether the observed behavioral and neural changes persist beyond the immediate intervention phase. This longitudinal component is designed to assess the sustainability of tDCS effects and explore delayed neuroplastic adaptations.

### 2.6 Data collection and analysis

Behavioral data will be collected at multiple time points to evaluate the effects of tDCS on cognitive function rehabilitation in individuals with schizophrenia. Cognitive performance will be assessed using the CANTAB battery during V1 and V2 to analyze short-term changes (Fig 2). A follow-up assessment conducted eight weeks after the completion of the intervention (V3) will allow for the evaluation of long-term or sustained effects (Fig 2).

Resting 20-minute EEG data will be recorded during V1, V2, and V3 (Fig 2) using a 21-channel dry electrode cap positioned according to the international 10–20 system. EEG processing pipeline will include filtering, referencing, semi-automatic analysis of bad channels, removal of artifacts, fixed-time epoching and further analysis. EEG recordings will be processed using the NeuroAnalyzer toolbox (https://neuroanalyzer.org). The authors will perform an exploratory analysis in the frequency domain (spectral analysis), examine various connectivity parameters (e.g., changes in coherence), entropy, mutual information, Higuchi Fractal Dimension and others.

At the same time points (V1, V2, V3), the Positive and Negative Syndrome Scale (PANSS) and the Clinical Global Impression – Severity scale (CGI-S) will be used.

For all tDCS sessions, data will be collected on the most frequent side effects: 1) itching/tingling, 2) pain, 3) burning sensation, 4) warmth sensation, 5) metallic taste, 6) tiredness, 7) headache and 8) other side effects. For each side effect, subjective assessment on the severity of symptoms was obtained from each patient (immediately after the stimulation was finished), using a scale from 0 (complete lack of), 1 (very mild), 2 (mild), 3 (moderate), 4 (severe) to 5 (extremely severe). The protocol also included details on the onset (from the beginning of the current session, during the session or at its end), duration (only at the beginning, lasting until the middle of a session or lasting by the end of stimulation), subjective level of distress (mild, moderate or significant) and location (both anode and cathode, only anode, only cathode or generalized). If there were any adverse effects observed by the doctor performing tDCS sessions, it was also reported. In all the observed cases, it was redness of the skin underneath the electrodes. Its severity was assessed on a scale from 1 to 5 (1-very mild, 2-mild, 3-moderate, 4 – severe, 5-extremely severe). The tDCS device will be administered by a therapist with specialized training and experience in its application. Consequently, it is expected that participants will not experience any significant health risks or adverse events. Moreover, after each procedure, patients will be asked to guess whether they have received real or sham stimulation in order to evaluate the blinding.

Histograms, QQ-plots and the Shapiro-Wilk test will be used to analyze whether the data fall within the normal range. If the quantitative data conform to a normal distribution, they will be expressed as the mean and standard deviation (SD); if not, they will be displayed as median values and interquartile range (IQR). Categorical data will be presented as counts with percentages. Statistical tests, appropriately selected for the nature of the data and the objectives of the study, will be applied to make the analyses.

Data will not be collected from participants who discontinue or deviate from the intervention protocol; therefore, no missing data are expected in the primary analysis. Analyses will be performed according to the per-protocol (PP) principle, including only participants who strictly adhered to the study protocol and completed at least two visits. If data from visit 3 are missing, the most recent available data from a prior visit will be used for the analysis.

To evaluate between-group and within-group differences over time for both primary and secondary outcomes, the authors will appropriately apply repeated measures ANOVA. To account for multiple comparisons across multiple time points, p-values will be adjusted using the Bonferroni correction method. The threshold of statistical significance will be set at < 0.05.

No interim analyses or early stopping of the trial are planned. Any decision to terminate the study will be made by the principal investigator in consultation with the ethics committee.

## 3 Discussion

In our study, we will use the RehaCom system, which is a computer-assisted therapy aimed at improving cognitive functioning (Garcia-Fernandez, 2019). Its effectiveness in enhancing cognitive outcomes has been demonstrated in randomized controlled trials among patients with schizophrenia in remission (d’Amato et al., 2011). In light of the above, neuroplasticity may be an important mechanism underlying effective intervention methods.

tDCS induces polarity-dependent changes in cortical excitability both during and after stimulation. Anodal stimulation increases excitability, whereas cathodal stimulation decreases it (Nitsche et al., 2003). Animal studies have demonstrated that the effects of anodal tDCS are due to neuronal depolarization, whereas cathodal stimulation leads to hyperpolarization of cortical neurons (Nitsche et al., 2003). Importantly, when applied for ten minutes, tDCS has been shown to induce changes in cortical excitability that persist for more than an hour after the stimulation ends (Nitsche and Paulus, 2001). The ability to induce long-lasting, non-invasive, painless, and reversible modulations of excitability makes tDCS a potentially valuable tool for modulating neuroplasticity (Nitsche and Paulus, 2001). Its therapeutic effect is thought to result from its influence on cellular and molecular mechanisms involved in long-term potentiation (LTP) (Stagg et al., 2009). LTP, in particular, promotes increased synthesis of various proteins, such as neurotransmitter synthases, receptors, ion channels, and intracellular signaling proteins, thereby enhancing neurotransmission efficiency within the cortical circuits (Yamada, 2021a). Consequently, if applied before the cognitive training, tDCS seems to be a very promising intervention aimed at improving the efficacy of cognitive rehabilitation.

Currently, there are few studies evaluating the combined effect of cognitive training and transcranial direct current stimulation in patients with schizophrenia, and most of them involved small groups, which limits the representativeness of the results.

In one study conducted by Padinjareveettil et al. (2015), two patients received CR intervention combined with tDCS, which resulted in cognitive improvement that lasted throughout a one-month follow-up. The intervention lasted four weeks and included five 45-minute auditory training sessions per week, with active tDCS applied simultaneously with CR during three sessions per week. The anode was placed over the left dorsolateral prefrontal cortex, and the cathode over the right orbitotemporal area; stimulation lasted 20 minutes at an intensity of 2 mA (Jahshan, 2017).

In another pilot, randomized, sham-controlled, single-blind proof-of-concept study, ten patients participated in three working memory training sessions per week, each lasting one hour, for 16 weeks. Beginning in the third week, tDCS or sham stimulation was applied for the first 20 minutes of two weekly sessions, for a total of 28 sessions. Stimulation parameters included 1 mA, with the anode placed over the left DLPFC and the cathode over the contralateral supraorbital area. Training tasks included, among others, PSS CogRehab, Captain’s Log (BrainTrain), and N-back tasks with words and images. The results suggested that combining tDCS with working memory training led to a greater improvement in cognitive function compared to training alone (Nienow TM, 2016).

In a four-week, double-blind, sham-controlled RCT involving 12 patients (eight with schizophrenia and four with schizoaffective disorder), participants were randomly assigned to the active tDCS group or the sham group. In the active group, asymmetric stimulation was applied: anodal 2 mA over the right DLPFC and cathodal over the left temporoparietal junction for 20 minutes, simultaneously with performing a computerized 2-back spatial working memory task. After two weeks of active stimulation, improvements were observed in untrained working memory tests as well as verbal fluency, with a significant verbal fluency enhancement maintained also after four weeks of treatment. In summary, a four-week anodal tDCS over the DLPFC appears to induce lasting beneficial cognitive effects in patients with schizophrenia, which also generalize to other cognitive domains dependent on the frontal lobes (Weickert, 2019).

In one of the largest double-blind, sham-controlled studies conducted to date, involving 49 patients, participants received CR (working memory training and implicit learning tasks) combined with either active (n = 24) or sham (n = 25) tDCS. The intervention included four days of cognitive training (days 1, 2, 14, and 56), with tDCS applied simultaneously with CR on days 1 and 14. Active stimulation was delivered continuously for 30 minutes at 2 mA; the anode was placed over the left DLPFC and the cathode over the right supraorbital area. The results showed significantly better working memory performance in the CR + active tDCS group both in next-day measurement and long-term follow-up, with no acute enhancement effect during stimulation. However, it is worth noting that tDCS was applied in two selected sessions only (Orlov, 2017).

In a more recent double-blind, placebo-controlled study, 28 patients with schizophrenia or schizoaffective disorder underwent ten-day adaptive working memory training (aWMT) lasting 22 minutes, combined with simultaneous anodal tDCS over the right DLPFC; the cathode was placed over the left deltoid muscle. The current intensity was 2 mA, and the total duration of stimulation was 25.5 minutes. The study provided preliminary evidence for effective enhancement of cognitive training with tDCS. Improvements in working memory and learning were partially maintained at follow-up assessments, and importantly, clinically significant transfer effects to untrained cognitive domains were also observed (Schwippel, 2025).

Reviews of the literature (e.g., Jashan et al.) indicate that tDCS is a promising tool for enhancing the effects of CR in schizophrenia, however, they also emphasize the need for further research to clearly determine the efficacy of the combined intervention. Additionally, they highlight the lack of standardization of tDCS parameters, including the appropriate dose (number of sessions per day or week), electrode placement, current amplitude, and selection of training tasks during stimulation (Jahshan, 2017).

Nevertheless, not all studies confirm the beneficial effect of combining tDCS with CR. In a randomized, double-blind study conducted by Shiozawa et al. (2016), ten patients were randomly assigned to active or sham tDCS groups (ten sessions over five days, 2 mA for 20 minutes, with the cathode over the right DLPFC, and the anode over the left DLPFC), combined with N-back training and sequence learning tasks. The results did not show significant cognitive improvement in patients with schizophrenia (Shiozawa P, 2016).

In conclusion, existing data suggest that combining CR with tDCS may lead to improvements in cognitive function in patients with schizophrenia. However, the absence of tDCS in all cognitive training sessions and the small sample sizes limit the ability to draw definitive conclusions. Larger, standardized, randomized clinical trials are necessary to confirm efficacy and determine optimal tDCS parameters in combination with CR in patients with schizophrenia. The authors think that the information presented in the present study expands knowledge on the efficacy of CR combined with tDCS and may contribute to improved treatment outcomes of cognitive impairment not only in schizophrenia but also other neuropsychiatric disorders.

## 4 Ethics and Dissemination

### 4.1 Ethics

Data management will be conducted in accordance with ethical data protection standards. All data will be stored on a secure, encrypted server with access limited to authorized personnel. To ensure patient confidentiality, pseudonymization will be employed by substituting real patient names with unique identification numbers assigned at data entry. Data entry will follow standardized procedures with double-entry verification and automated range checks to ensure data quality.

A Data Monitoring Committee (DMC) will not be established for this study, as the intervention (tDCS combined with RehaCom cognitive training) is considered low risk. Participant safety and adverse events will be regularly monitored by the research team throughout the study.

No specific ancillary or post-trial care is planned, as the intervention is considered low-risk. Participants who experience any adverse effects related to the study will receive appropriate medical care, and any compensation will be provided in accordance with local regulations.

### 4.2 Dissemination

The results of this study will be disseminated through publication in a peer-reviewed journal and presentation at scientific conferences. The study findings will also be reported in the clinical trial registry. Participants will be able to obtain information about the study results upon request.

## Data Availability

No datasets were generated or analysed during the current study. All relevant data from this study will be made available upon study completion.

## Acknowledgments

None

